## Supplementary Figures for "Rare variant associations with birth weight identify genes involved in adipose tissue regulation, placental function and insulin-like growth factor signalling"

**Supplementary Figure 1** | Comparison of fetal and maternal exome-wide significant gene burden associations for birth weight, as identified by the two centres. All effect estimates and accompanying 95% CIs are presented in phenotypic standard deviations (SDs). Relevant data is included in Supplementary Table 1.

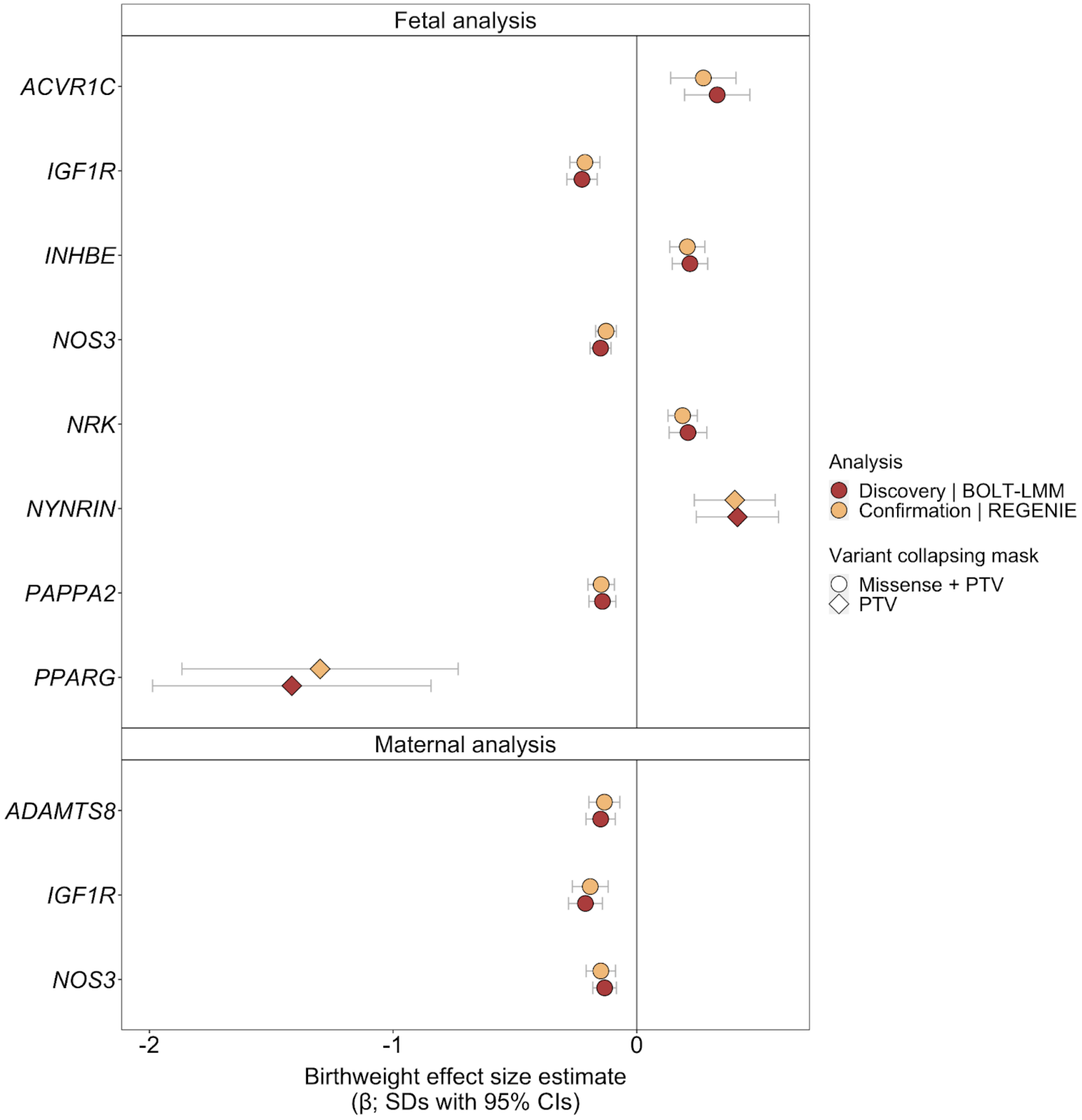

**Supplementary Figure 2** | Fetal variant-level associations between *IGF1R* (a), *NOS3* (b), *NRK* (c), *NYNRIN* (d), *PAPPA2* (e) and birth weight in BOLT-LMM. Included variants had a minor allele frequency (MAF) <0.1% and were annotated to be high confidence protein truncating variants (PTV) or PTVs plus missense variants with a CADD score ≥25 (Missense + PTV). Each variant is presented as an individual line extending to its association *p*-value (-$\log_{10}$) in the direction indicating the direction of effect on birth weight in carriers of the alternate allele. The point size indicates the number of carriers in each variant (i.e. allele count, AC). Plots depicting variants in *ACVR1C*, *INHBE* and *PPARG* can be found in Figures 3 and 4.

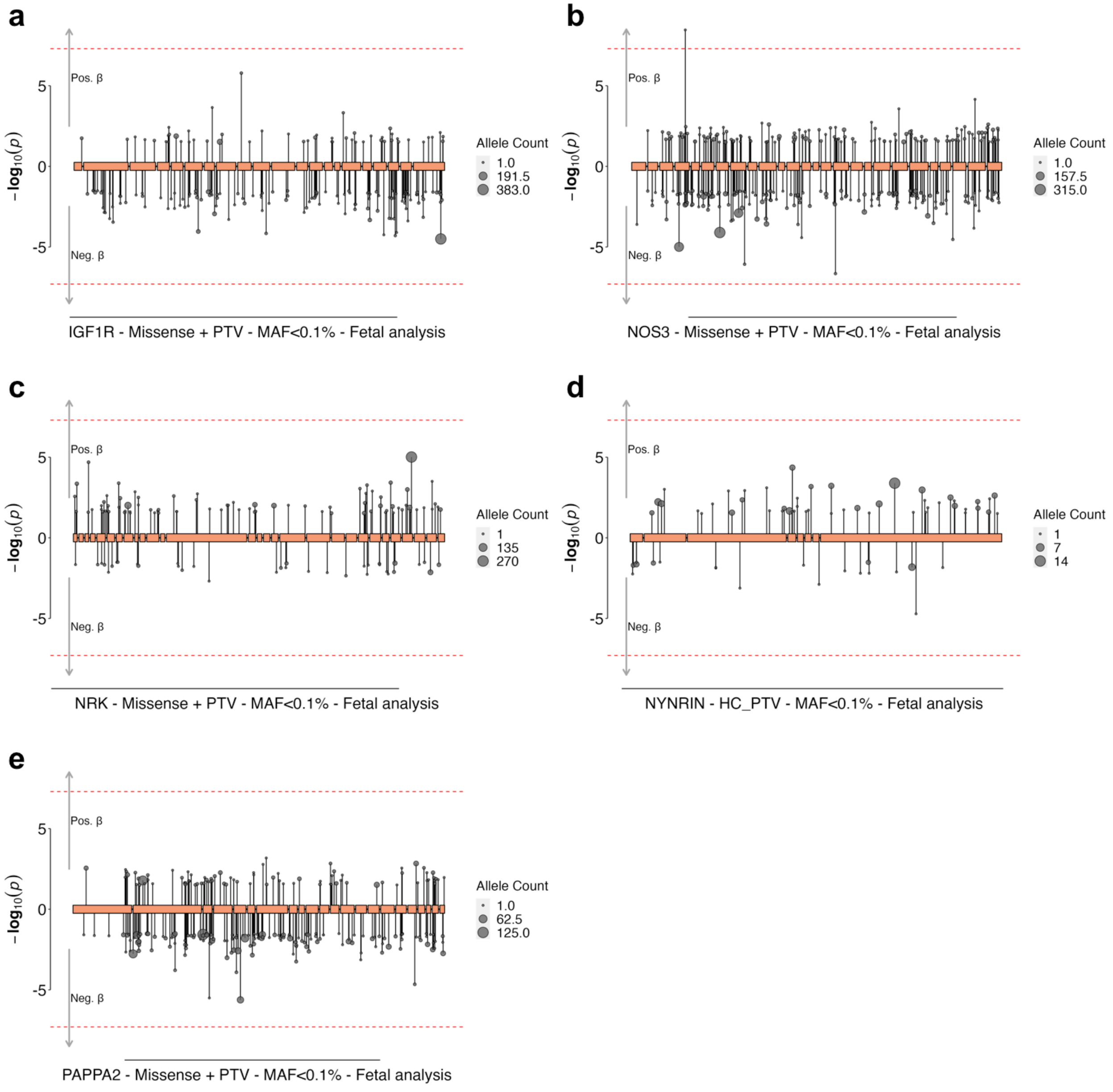

**Supplementary Figure 3** | Fetal variant-level associations between *ACVR1C* (a), *IGF1R* (b), *INHBE* (c), *NOS3* (d), *NRK* (e), *NYNRIN* (f), *PAPPA2* (g), and *PPARG* (h) and birth weight in REGENIE. Included variants had a minor allele frequency (MAF) <0.1% and were annotated to be high confidence protein truncating variants (PTV) or PTVs plus missense variants with a CADD score ≥25 (Missense + PTV). Each variant is presented as an individual line extending to its association *p*-value (-$\log_{10}$) in the direction indicating the direction of effect on birth weight in carriers of the alternate allele. The point size indicates the number of carriers in each variant (i.e. allele count, AC).

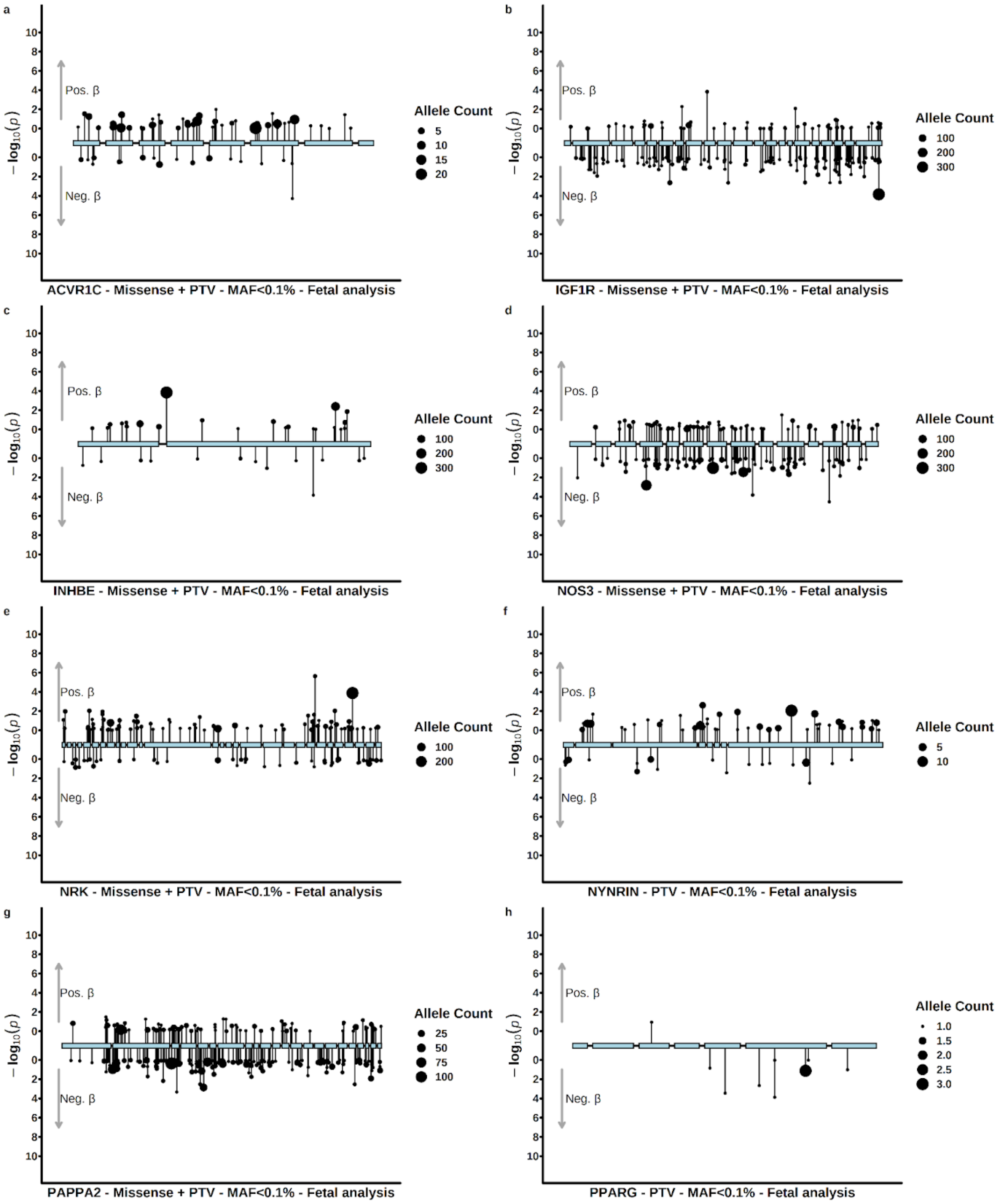

**Supplementary Figure 4** | Maternal variant-level associations between *ADAMTS8* (a), *IGF1R* (b), *NOS3* (c) and birth weight in BOLT-LMM. Included variants had a minor allele frequency (MAF) <0.1% and were annotated to be high confidence protein truncating variants (PTV) or PTVs plus missense variants with a CADD score ≥25 (Missense + PTV). Each variant is presented as an individual line extending to its association *p*-value ($-\log_{10}$) in the direction indicating the direction of effect on birth weight in carriers of the alternate allele. The point size indicates the number of carriers in each variant (i.e. allele count, AC).

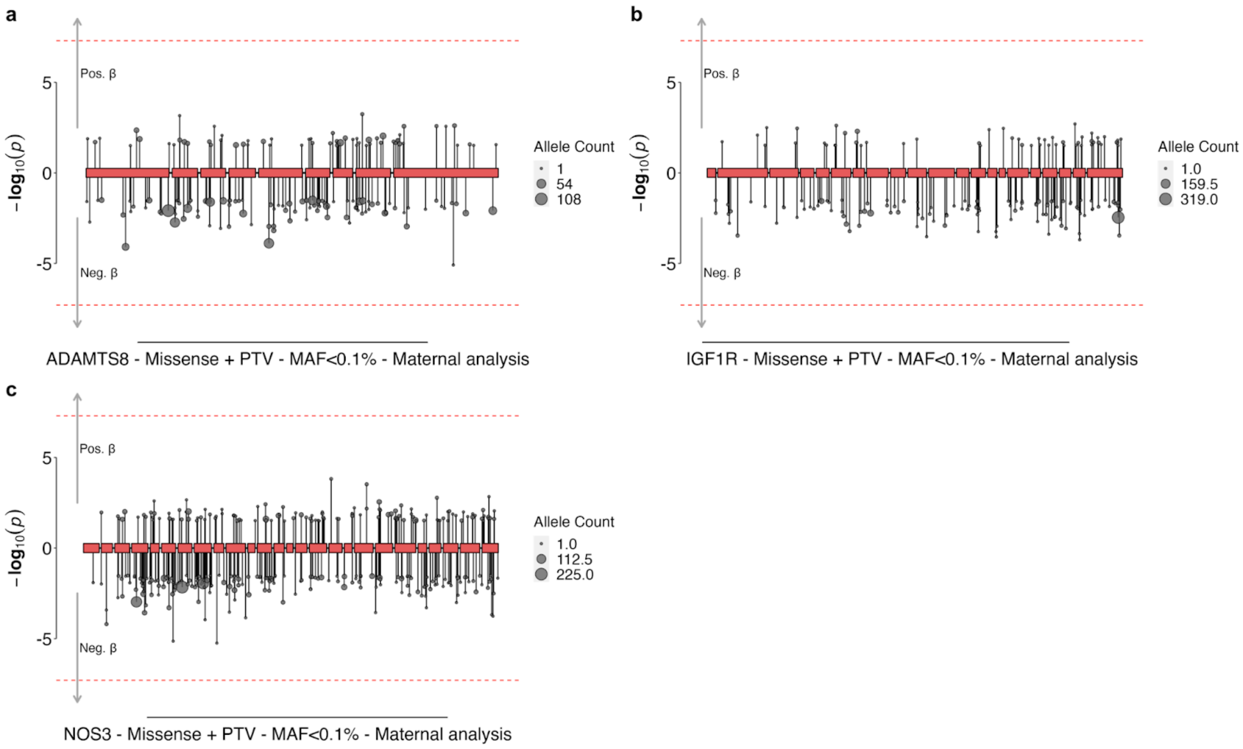

**Supplementary Figure 5** | Maternal variant-level associations between *ADAMTS8* (a), *IGF1R* (b), *NOS3* (c) and birth weight in REGENIE. Included variants had a minor allele frequency (MAF) <0.1% and were annotated to be high confidence protein truncating variants (PTV) or PTVs plus missense variants with a CADD score ≥25 (Missense + PTV). Each variant is presented as an individual line extending to its association *p*-value (-log$_{10}$) in the direction indicating the direction of effect on birth weight in carriers of the alternate allele. The point size indicates the number of carriers in each variant (i.e. allele count, AC).

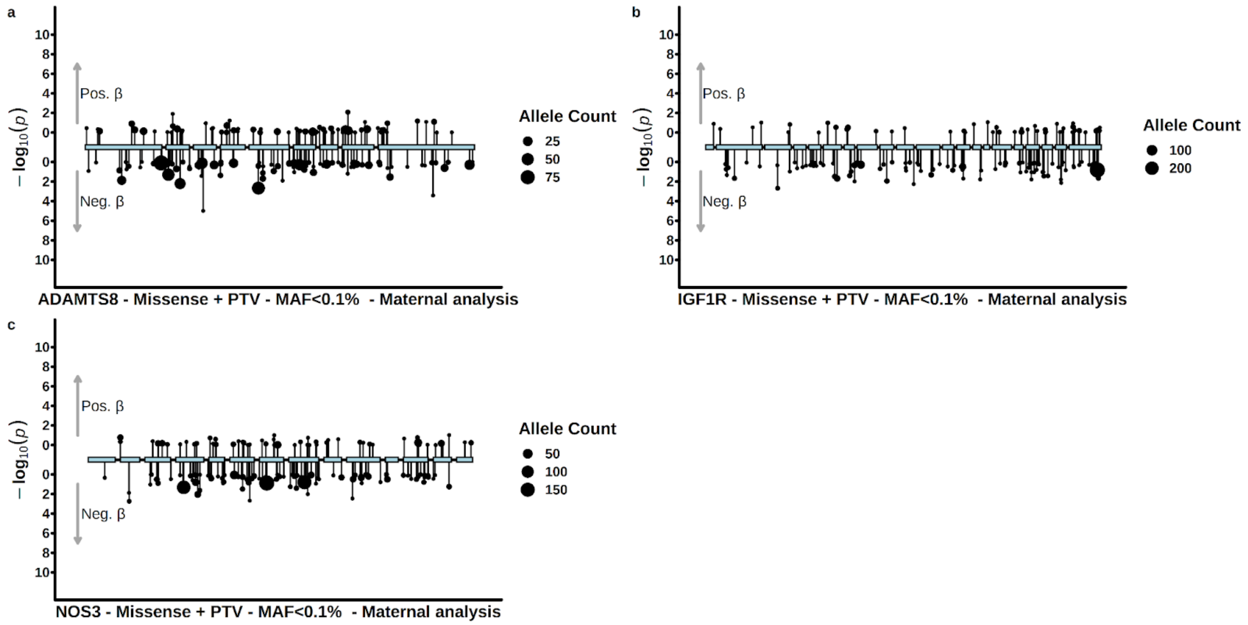

**Supplementary Figure 6** | Replication of the fetal and maternal exome-wide significant gene burden associations for birth weight in an independent Icelandic cohort. All effect estimates and accompanying 95% CIs are presented in phenotypic standard deviations (SDs). Relevant data is included in Supplementary Table 4.

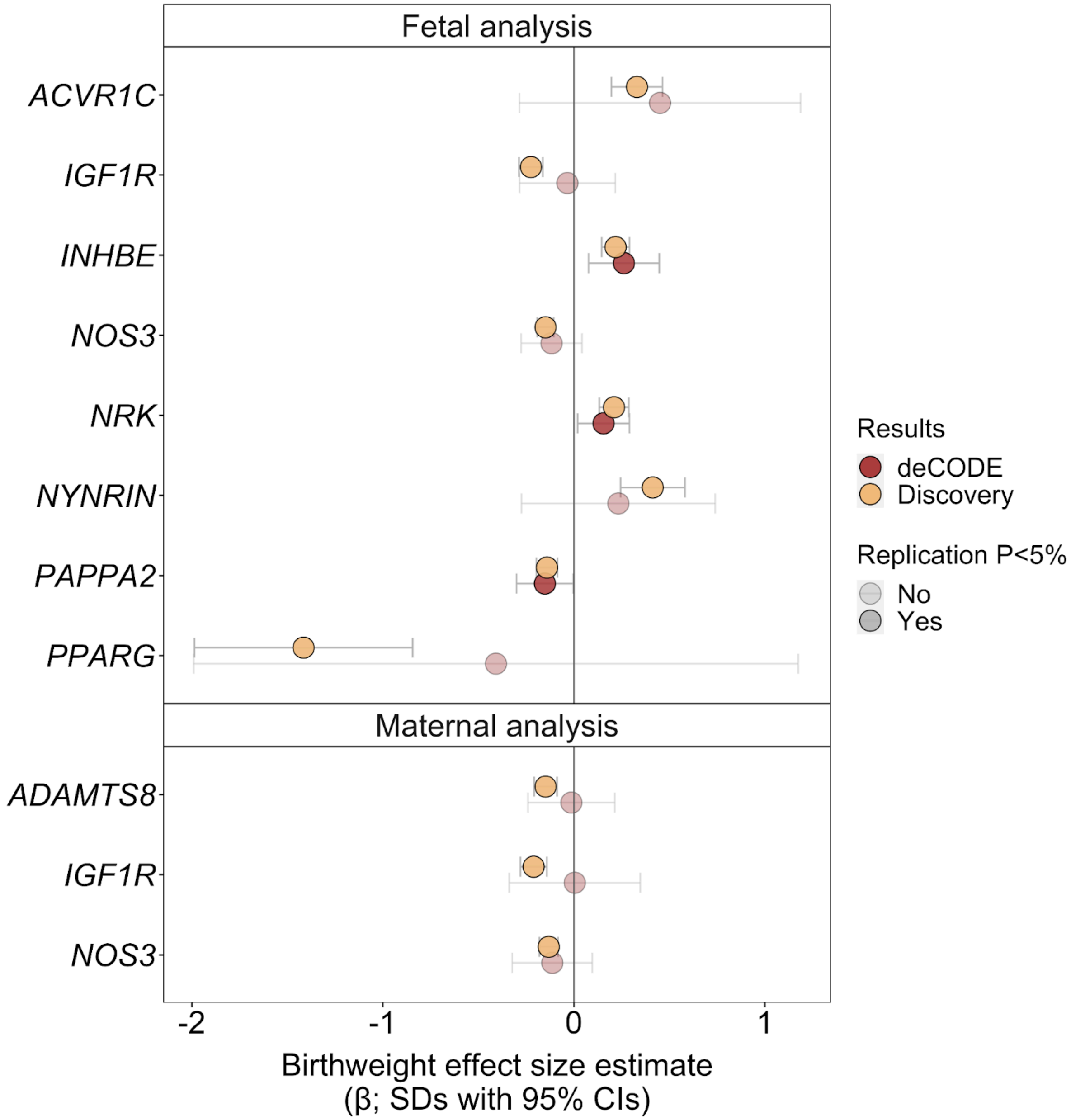

**Supplementary Figure 7** | Locus zoom plots of fetal GWAS loci for birth weight from Juliusdottir *et al.* are shown surrounding *ACVR1C* (a), *IGF1R* (b), *PAPPA2* (c), as well as the maternal birth weight locus around *NOS3* (d). Relevant data is included in Supplementary Table 5.

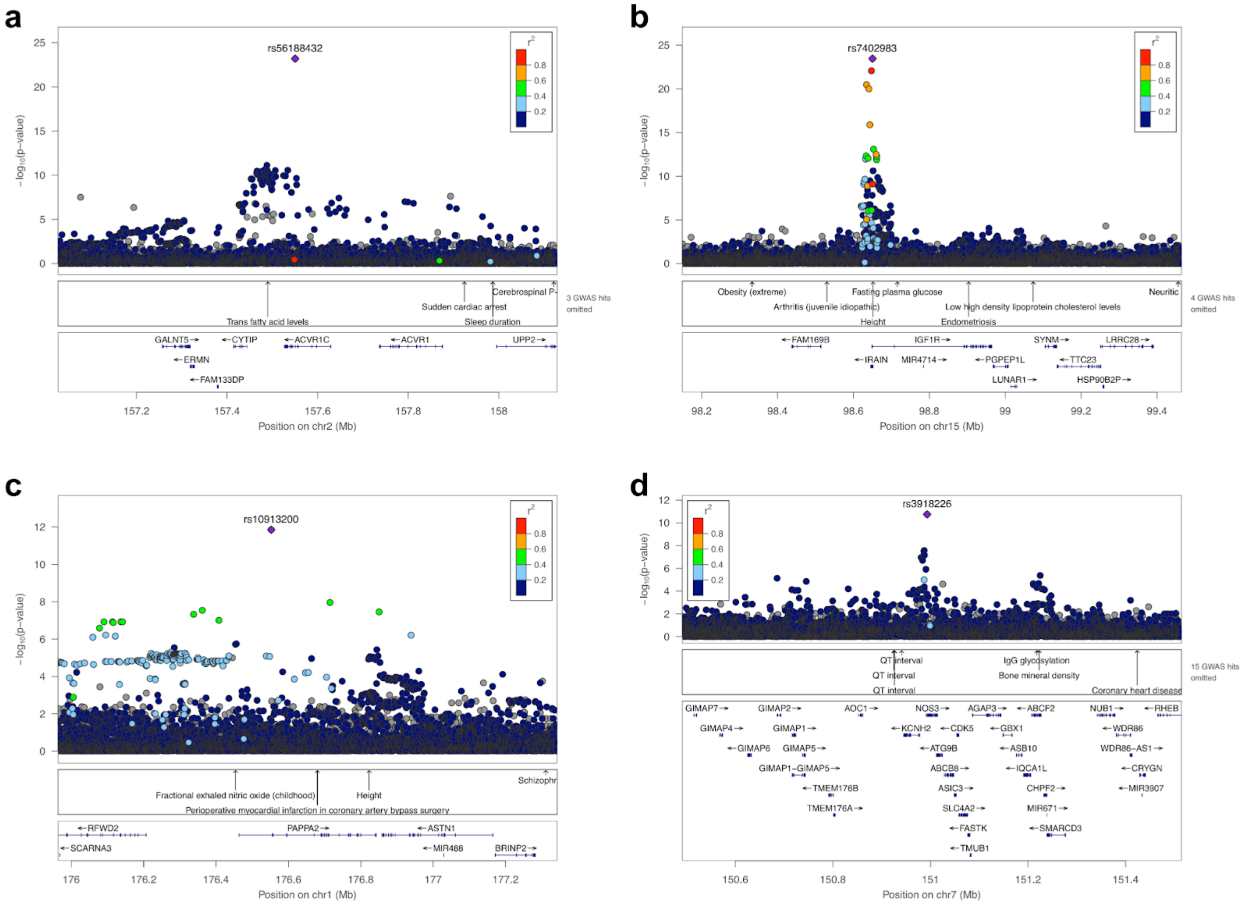

**Supplementary Figure 8** | Comparison of associations between known pathogenic variants in *GCK* and birth weight with the REGENIE burden test results. Beta values are in birth weight SD units; error bars represent 95% confidence intervals. Relevant data is included in Supplementary Table 12.

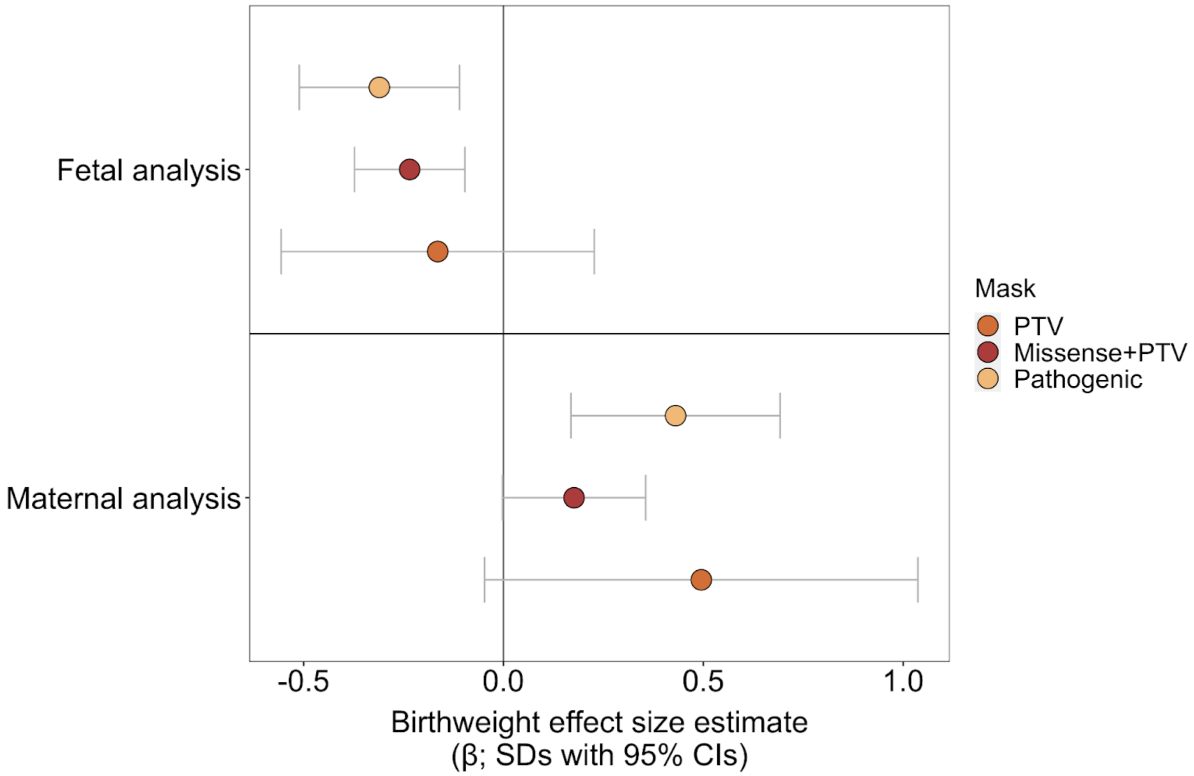
